# Humoral Immunity to SARS-CoV-2 and Inferred Protection from Infection in a French Longitudinal Community Cohort

**DOI:** 10.1101/2022.05.23.22275460

**Authors:** Tom Woudenberg, Laurie Pinaud, Laura Garcia, Laura Tondeur, Stéphane Pelleau, Alix De Thoisy, Françoise Donnadieu, Marija Backovic, Mikaël Attia, Nathanael Hozé, Cécile Duru, Aymar Davy Koffi, Sandrine Castelain, Marie-Noelle Ungeheuer, Sandrine Fernandes Pellerin, Delphine Planas, Timothée Bruel, Simon Cauchemez, Olivier Schwartz, Arnaud Fontanet, Michael White

**Author notes:** **Correspondence** Dr Michael White, Infectious Disease Epidemiology and Analytics G5 Unit, Department of Global Health, Institut Pasteur, Université Paris-Cité, Prof Arnaud Fontanet, Emerging diseases epidemiology unit, Institut Pasteur, Université Paris-Cité, Paris, France.

## Abstract

Population-level immunity to SARS-CoV-2 is growing through vaccination as well as ongoing circulation. Given waning immunity and emergence of new variants, it is important to dynamically determine the risk of re-infection in the population. For estimating immune protection, neutralization titers are most informative, but these assays are difficult to conduct at a population level. Measurement of antibody levels can be implemented at high throughput, but has not been robustly validated as a correlate of protection. Here, we have developed a method that predicts neutralization and protection based on variant-specific antibody measurements to SARS-CoV-2 antigens. This approach allowed us to estimate population-immunity in a longitudinal cohort from France followed for up to 2 years. Participants with a single vaccination or immunity caused by infection only are especially vulnerable to COVID-19 or hospitalization due to SARS-CoV-2. While the median reduced risk to COVID-19 in participants with 3 vaccinations was 96%, the median reduced risk among participants with infection-acquired immunity only was 42%. The results presented here are consistent with data from vaccine-effectiveness studies indicating robustness of our approach. Our multiplex serological assay can be readily optimized and employed to study any new variant and provides a framework for development of an assay that would include protection estimates.

## Introduction

The coronavirus disease 2019 (COVID-19) pandemic has resulted in significant morbidity and mortality worldwide with an estimated 18.2 million deaths (1), and 3.8 billion infections and reinfections as of November 2021, infecting 43.9% of the world’s population (2). In addition to infection-acquired immunity, a large share of the world has been vaccinated. By December 2021, 65% of the US population was vaccinated at least once (3), and in the European Union, 75% had at least one vaccination, with wide variation from 30% to over 90% between countries (4).

Population-level immunity can be measured with serology-based assays. Sero-prevalence is determined by measuring the presence of antibodies to a particular protein of SARS-CoV-2, often either whole Spike, a smaller component of Spike such as its receptor-binding domain (RBD), or Nucleocapsid protein (NP). The presence of antibodies is associated with protection against infection, but is not always predictive of protection (5). Unlike most serological assays, a neutralization assay measures antibodies that can block viral replication and infection of cells. These so-called neutralizing antibodies are more likely to provide protection as they possess a direct antiviral activity. Despite strong individual correlations between antibodies levels and neutralization activity, individuals with similar IgG levels following vaccination were regularly observed to have substantially varying neutralization titers (6).

By combining neutralization titers from immunological trials and corresponding efficacy estimates from vaccine trials, it was shown that neutralization titers correlate very well with protection against symptomatic infection and hospitalization (7-9), with higher neutralization titers associated with higher vaccine efficacy. With the advent of new variants that partly escape immunity, these correlates had to be updated. Relative to neutralization titers to ancestral strains, neutralization titers reduced 4-fold to Delta, and 1.6 fold to Alpha variants. (10, 11). For these variants, the variant-specific neutralization titers remained strongly correlated with protection outcomes and aligned well with new vaccine effectiveness estimates (12). However, Omicron caused a further reduction in neutralization. Relative to Delta, neutralization titers of sera with high antibody levels reduced 6-23 fold against Omicron (13).

Assessment of population-level immunity can provide critical information in the response to the SARS-CoV-2 pandemic, for example by identifying vulnerable subgroups in need of control measures such as boosters. Neutralization titers are not optimal for population-level surveillance because they are too time-consuming to process large sample numbers, and require at least cell-culture equipment, which are not present in many laboratories. Widespread measurement of population-level immunity requires high-throughput assays that can be more easily implemented in diagnostic laboratories.

Here, we develop a multiplex serological assay measuring the binding of antibodies of different isotypes to a variety of SARS-CoV-2 antigens as well as the strength of these interactions. The correlation between these measurements and neutralization activities against multiple variants was used to develop a prediction model for serum samples. From the predicted neutralization titers, we use previously developed models (7, 12) to translate these titers into individual-level protection estimates. We applied this method to a longitudinal cohort study of approximately 900 individuals followed for up to two years.

## Methods

### Samples

#### Viral neutralization studies

To correlate antibody measurements with neutralization titers, we collected 304 serum samples from individuals with either vaccine-induced or infection-acquired immunity to SARS-CoV-2. These individuals were enrolled in two different clinical cohort studies, described elsewhere in more detail (10, 11). Individuals who participated in the Orleans cohort were either convalescent or vaccinated. Researchers aimed to describe the persistence of specific and neutralization antibodies over a 24-month period. The second clinical cohort study, the Strasbourg cohort, included convalescent individuals only. Serological status of these individuals were described at three or six months after symptom onset.

#### COVID-Oise cohort

One of the first clusters of COVID-19 in France was detected in the town of Crépy-en-Valois in Oise Department. In winter 2020, scientists at Institut Pasteur initiated a longitudinal cohort study, named the COVID-Oise cohort. Participants comprise a wide age range starting from 5 years-old, up to nursing home residents. Participants have been invited four times to collect epidemiological data and biological specimen. Data and samples from the three first collection sessions are used in the current manuscript. Many of the COVID-Oise participants also participated in earlier studies that took place in spring 2020. Collected data at that time revealed high attack rates in a high school prior to the first reported case of SARS-CoV-2 in the community (14). Collected data and biological specimen from these earlier studies were integrated to the analyses in the current manuscript.

### Serological assays

#### Luminex assay

In a 96 well, non-binding microtiter plate 50 μL of protein-conjugated magnetic beads (250/region/well) and 50 μL of serum diluted 1/100 for IgG assay or 1/200 for IgA assay were mixed and incubated for 30 min at room temperature on a plate shaker. All dilutions were made in phosphate buffered saline containing 1% bovine serum albumin and 0.05% (v/v) Tween-20 (denoted as PBT). Following incubation, the magnetic beads were separated using magnetic plate separator (Luminex^®^) for 60 seconds and washed thrice with 100 μl PBT. The washed magnetic beads were incubated for 15 minutes with detector secondary antibody at room temperature on a plate shaker, washed thrice with 100 μl PBT and finally resuspended in 100 μL of PBT. R-Phycoerythrin-(R-PE) conjugated goat or donkey anti-human IgG antibody was used as detector antibody at 1/120 dilution and goat anti-human IgA at 1/200. A positive control pool of serum at two-fold serial dilutions from 1:50 to 1:102,400 was included on each 96 well plate. Plates were read using a Luminex^®^ MAGPIX^®^ system, which provides a reading of median fluorescence intensity (MFI).

A previously described 9-plex bead-based assay was extended to detect antibodies to 30 antigens in 1 μL serum or plasma samples (15). This assay allowed simultaneous detection of antibodies to 30 antigens, including stabilized trimeric Spike ectodomain (16), RBD, Membrane protein (M), Membrane Envelope protein (E), Nucleocapsid protein (NP), and a Membrane-Envelope fusion protein (ME). The trimeric Spike ectodomains and RBD antigens were produced as recombinant proteins for four SARS-CoV-2 variants, namely of the ancestral lineage, Alpha, Beta, and Delta variants. In addition, we included 8 antigens of 4 seasonal coronaviruses (Spike ectodomain and NP of NL63, 229E, HKU1, OC43). ME and Spike Sub-unit-2 (S2) SARS-CoV-2 antigens were purchased from Native Antigen (Oxford, United Kingdom) and all other antigens were produced as recombinant proteins at Institut Pasteur. The mass of proteins coupled on beads was optimized to generate a log-linear standard curve with a pool of 27 positive sera prepared from patients with reverse-transcription quantitative PCR–confirmed SARS-CoV-2 (15). We measured the levels of immunoglobulin G (IgG) and immunoglobulin A (IgA) of each sample in two separate assays. Plates were read using a Luminex MAGPIX system and the median fluorescence intensity (MFI) was used for analysis. A 5-parameter logistic curve was used to convert MFI to relative antibody units (RAU), relative to the standard curve performed on the same plate to account for inter-assay variation.

In addition to the measurement of the presence of antibodies to antigens, we also measured the strength of antibody (Ab) binding with an avidity assay (Garcia et al, submitted to Viruses, 2022). The protocol for the avidity assay was similar to the serological assay with the inclusion of an additional step. After incubation of beads and serum samples, the complex beads-Ab were washed and then incubated for 5 minutes with 100μl of urea 6 M diluted in water, or water alone. After these 5 minutes and washing, 100μL of secondary antibodies conjugated to R-phycoerythrin (Jackson Immunoresearch) for detection of specific IgG, diluted at 1/100 was added for 15 minutes. To finish, after washing, plates were read using a Luminex^®^ MAGPIX^®^ system and the median fluorescence intensity (MFI) was used for analysis. The Avidity index (AI) was measured with AI = [MFI after treatment with 6M of Urea/ MFI without Urea] x 100. Avidity was only assayed for IgG.

#### S-Fuse neutralization assay

U2OS-ACE2 GFP1-10 or GFP 11 cells, also termed S-Fuse cells, become GFP+ when they are productively infected by SARS-CoV-2 (10, 17). Cells tested negative for mycoplasma. Cells were mixed (ratio 1:1) and plated at 8 × 103 per well in a μClear 96-well plate (Greiner Bio-One). The indicated SARS-CoV-2 strains were incubated with serially diluted monoclonal antibodies or sera for 15 min at room temperature and added to S-Fuse cells. The sera were heat-inactivated 30 min at 56⍰°C before use. Eighteen hours later, cells were fixed with 2% paraformaldehyde (PFA), washed and stained with Hoechst (dilution 1:1,000, Invitrogen). Images were acquired with an Opera Phenix high-content confocal microscope (PerkinElmer). The GFP area and the number of nuclei were quantified using Harmony software (PerkinElmer). The percentage of neutralization was calculated using the number of syncytia as value with the following formula: 100 × (1 – (value with serum − value in ‘non-infected’)/(value in ‘no serum’ − value in ‘non-infected’)). The neutralizing activity of each serum was expressed as the ED50 value. ED50 values (in μg ml−1 for monoclonal antibodies and in dilution values for sera) were calculated with a reconstructed curve using the percentage of the neutralization at the different concentrations.

#### Luciferase-Linked ImmunoSorbent Assay (LuLISA)

The LuLISA was used as a validation for the Luminex assay and for the determination of sero-positivity. Briefly, Nucleocapsid-specific IgG antibodies were assessed using an ELISA-based assays on sera incubated in antigen-coated wells. Antigens have been produced as follows. Full-length N protein from SARS-CoV-2 were produced with a (His)6 tag in the E. coli, purified on Ni-NTA affinity column, and then size-exclusion chromatography was performed. White 384-well plates with flat bottoms (Fluoronunc C384 Maxisorp, Nunc) were coated with 1 μg/mL of Nucleocapsid protein in PBS buffer, 50 μL/well for 3 h at room temperature, or overnight at 4°C. Wells were washed using a plate washer (Zoom, Berthold Technologies, Germany) two cycles of three times with 100 μL of PBS/Tween 20 0.1%. Sera were diluted 200 times in PBS, nonfat milk 1%, and Tween 20 0.1%. Note that 50 μL of serum dilutions were incubated for 1 h at room temperature in their respective wells. Wells were washed two cycles of three times with 100 μL of PBS/Tween 20 0.1%. The Anti-Fc IgG VHH (Fc1) was derived from an antibody from immunized alpaca and expressed as a tandem with an optimized catalytic domain nanoKAZ from Oplophorous gracilirostris luciferase. Purified Fc1-nanoKAZ 1 ng/mL (400 × 106 RLU·s–1·mL–1) in PBS, nonfat milk 1%, and Tween 20 0.1% was loaded (50 μL/well) and incubated for 30 min at room temperature. Wells were washed two cycles of three times with 100 μL of PBS/Tween 20 0.1% then 50 μL of the luciferin solution was added (Promega). Photons production was counted during 0.5 s per well and measured two times in a plate luminometer (Mithras2; Berthold, Wildbad, Germany).

### Statistical analyses

Relative antibody units, neutralization and protection estimates are visualized by vaccination and infection status. Vaccination status was self-reported. Infection status was determined through either a positive PCR or serology. Date of infection status was determined with the following strategy: (i) a positive PCR/Ag+ test confirmed by positive serology, (ii) clinical diagnosis by a medical doctor before the first positive serology, (iii) starting date of at least one self-reported symptom (fever, cough, dyspnea, agueusia/dysgueusia or anosmia/dysnomia) prior to a positive serology, or (iv) circulation of SARS-CoV-2 within a household or nursing home prior to a positive serology. If no infection date could be determined based on these four strategies, we used the mid-point between last negative serology and first positive serology. If no negative serology was available, the mid-point was calculated between January 1^st^ 2020 and first positive serology.

Serological classification of previous infection was dependent on vaccine status. For unvaccinated participants, we developed a random forest algorithm based on relative antibody units. This algorithm was trained on samples from both PCR-confirmed cases of COVID-19 and negative control samples, and calibrated to have 99% specificity (18). For vaccinated individuals, positivity was based on antibody levels to Nucleocapsid protein with both the LuLisA and the Luminex assay.

Protection in this study is defined to be against COVID-19 and severe COVID-19. These two terms have been used in different ways in the various efficacy studies (7). COVID-19 was usually a positive PCR combined with at least one or two typical symptoms, such as fever, cough, shortness of breath, chills, new or increased muscle pain, loss of taste or smell, sore throat, diarrhea, or vomiting. Severe Covid-19 was usually defined as confirmed COVID-19 with any of the following additional features: respiratory failure; evidence of shock; significant acute renal, hepatic, or neurologic dysfunction; admission to an intensive care unit; or death.

#### Estimation of neutralization titers

To establish a model to predict neutralization titers with relative antibody units from our multiplex assay, we tested samples using both the Luminex assay and the S-Fuse neutralization assay, and used the data to build random forest regression models. As we had SARS-CoV-2 variant-specific antigens for RBD and whole Spike for four variants, Ancestral, Alpha, Beta, and Delta, we developed four random forest regression models in parallel. For each random forest, the number of trees was set at 1000. For each tree, two-thirds of the observations were used. Predictions are derived from the average of a sample’s estimates in the remaining one third of the samples, the out-of-bag samples. Regressions were built in a step-wise manner. The first antigen in the regression was selected based on the importance of that antigen, measured by the mean decrease in accuracy on the out-of-bag samples. Subsequently, all other variables were added one by one to identify the most important antigen in the regression. The antigen associated with the lowest sum of residual sum of squares among the four different variant-specific random forest regression models was kept in the model. This process was repeated until no further decrease in the lowest residual sum of squares was observed.

#### Estimation of protection

Neutralization titers were normalized by the average neutralization activity in convalescent serum three weeks following symptom onset. Normalized neutralization titers were converted into protection estimates using models developed by Khoury et al and Cromer et al (7, 12). In short, the relationship between in vitro neutralization levels and the observed protection from SARS-CoV-2 infection was studied using immunogenicity data from phase 1 and 2 studies of seven vaccines and data on protection from corresponding phase 3 studies. A logistic model was used to describe the relationship. Subsequently, this model was extended with data from 24 studies on in-vitro neutralisation and clinical protection in order to incorporate the loss of neutralization to SARS-CoV-2 variants.

### Ethics

Collection of samples from the Orleans cohort had been approved by the Comité de Protection des Personnes Ile de France IV (NCT04750720). Collection of samples from the Strasbourg cohort was approved by Institutional Review Board of Strasbourg University Hospital (NCT04441684).

The study of the COVID-Oise cohort was registered with ClinicalTrials.gov (NCT04644159) and received ethical approval by the Comité de Protection des Personnes Nord Ouest IV. Several COVID-Oise participants participated in the CORSER studies in spring 2020, registered with ClinicalTrials.gov (NCT04325646) and approved by the Comité de Protection des Personnes Ile de France III.

For all studies, participants did not receive any compensation. Informed consent was obtained from all participants, and parents provided informed consent for any children under the age of 18 years. For the nursing home residents who did not have full capacity to sign legal documents, informed consent was obtained from their relatives.

## Results

### Estimation of neutralization titers

For 304 samples of the Viral neutralization studies, we measured neutralization titers to several SARS-CoV-2 variants (ancestral, Alpha, Beta, Delta) as well as IgG and IgA antibody levels and avidity to all SARS-CoV-2 and seasonal coronavirus antigens. Among these, 106 samples had immunity acquired through an infection and 198 had vaccine-acquired immunity of whom the majority were vaccinated twice with Pfizer (Table S1). We found a strong correlation between neutralization titers and relative antibody units, especially antibodies targeting Spike and RBD of isotype IgG (Figure 1a). Highest correlation is observed between IgG antibodies to Spike (r = 0.87) and neutralization activity. Visualizations for other variants are depicted in Figure S1.

**Figure 1.**
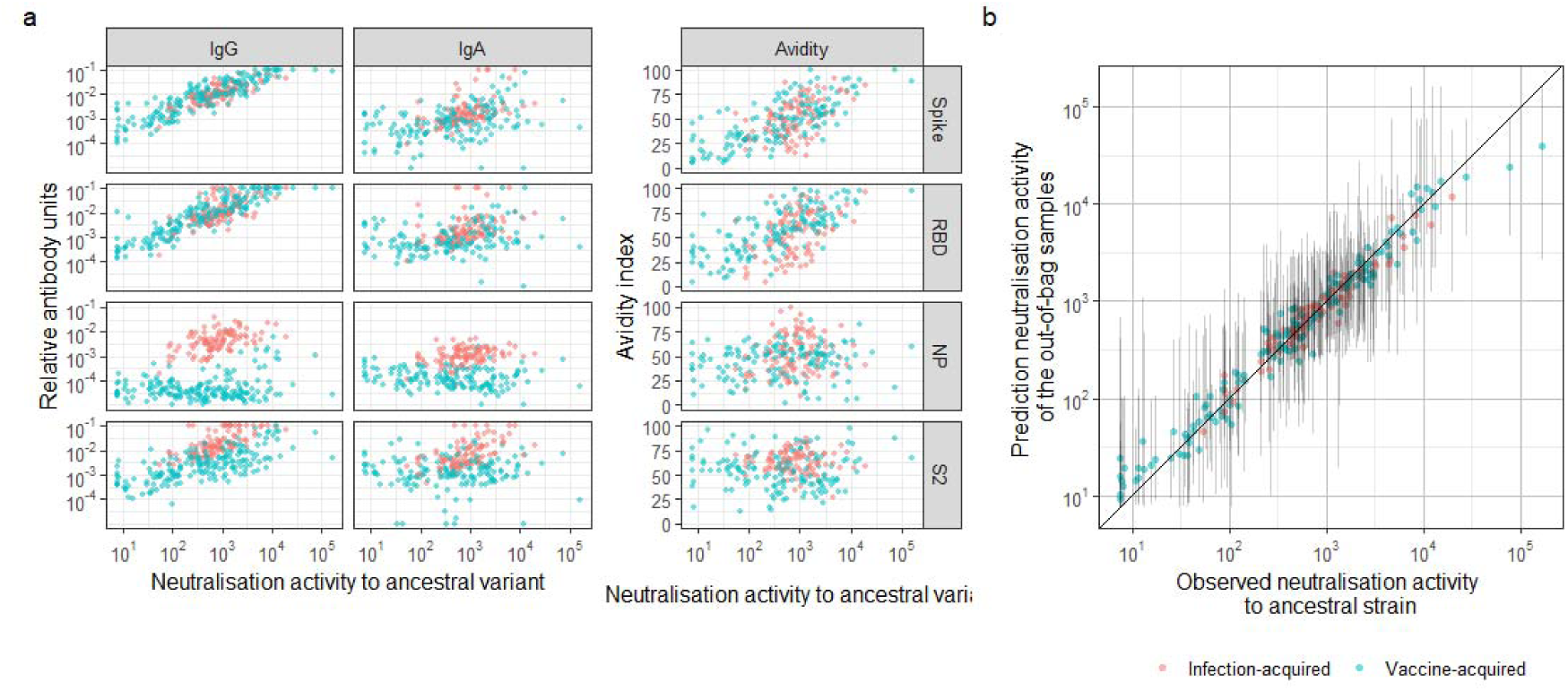
Establishment of a prediction model for neutralization activity based on measured antibody levels. a) Correlation between relative antibody units of Spike, RBD, NP and S2, for IgG, IgA and avidity and neutralizing activity to ancestral variant. b) Prediction of neutralization activity to the ancestral strain of SARS-CoV-2. The predictions were derived from a random forest regression model containing the following biomarkers: Spike IgG, RBD IgG, Spike IgG avidity, S2 IgG, RBD IgA and RBD IgG avidity.

With random forest regression models we found a strong association between RAU with variant-specific neutralization titers. Observed and predicted neutralization titers to ancestral lineage are shown in Figure 1b. Visualization of other variants are shown in Figure S2, where the development of the random forest regression models is shown as well. The final random forest regression comprised Spike IgG, RBD IgG, Spike IgG avidity, S2 IgG, RBD IgA, and RBD IgG avidity.

An alternative approach to predict neutralization titers to SARS-CoV-2 variants is to measure neutralization titers to the ancestral strain, and then adjust for the fold reduction in neutralization titers between variants. Taking into account the lower limit of detection, we described the relationships between neutralizing titers of the ancestral, Delta and Omicron strains using a censored linear regression model (Figure S3). We found that neutralization activity of samples against ancestral strain decreased on average by 62% (57% - 67%) against Delta. A further decrease was observed with Omicron, with a percentage decrease of 97.7% (97.1% – 98.3%) compared to the ancestral strain.

### COVID-Oise cohort

The COVID-Oise cohort was established in winter 2020 (session 1) and two follow-up sessions took place in spring 2021 and winter 2021 (sessions 2 and 3, Figure 2). 905 individuals have been enrolled during these three sessions. Several participants of the COVID-Oise cohort participated in earlier studies in spring 2020. Epidemiological data and serum samples were collected during each session. In total, 2.582 sera samples were collected. The initial studies held in spring 2020 led to the collection of a total of 487 sera samples for the participants who would later be enrolled in the COVID-Oise study. Three COVID-Oise sessions took place in winter 2020, spring 2021 and winter 2021. These sessions led to the collection of sera from 725, 750 and 620 participants, respectively. A detailed visualization of vaccinations, infections, and participation rates can be seen in Figure S4. Among participants who participated in April 2021 (session 2), 25% had at least one vaccination. Vaccination coverage of at least one vaccination increased to 87% in December 2021 (session 3).

**Figure 2.**
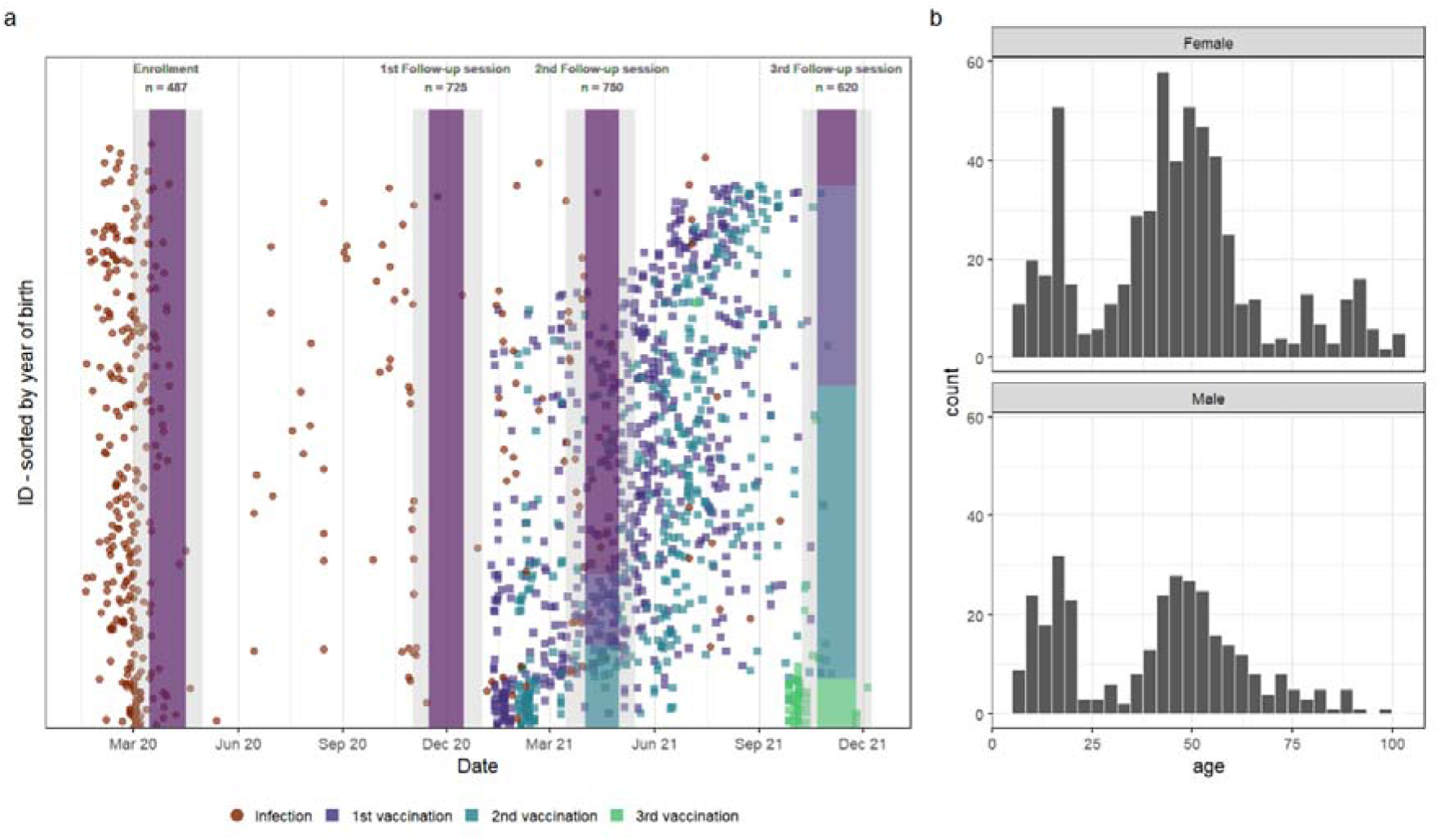
Overview of the study design and study population. (a) Analyses were based on four sessions of data and sera collection. Vaccination coverage is shown by the four stacked bar plots. Individual vaccination dates are shown in squares, color defines first, second and third dose. (b) Distribution of the study population by age and gender.

All samples were analyzed with the 30-plex serological assay, providing read outs for IgG, IgA and avidity. Among unvaccinated individuals, a clear distinction in the distribution of antibody levels to Spike, RBD, NP and S2 (Figure 3) was observed. In April 2020, 36% of all samples tested positive, which increased to 37% in November 2020, to 44% in April 2021 and 47% in November 2021. Antibody levels to IgA and avidity measurements by session are shown in Figures S5-8. A comparison of the measured Spike IgG and Nucleocapsid IgG between the Luminex multiplex assay and LuLISA showed a strong correlation as has been observed before (19) (Figure S9).

**Figure 3.**
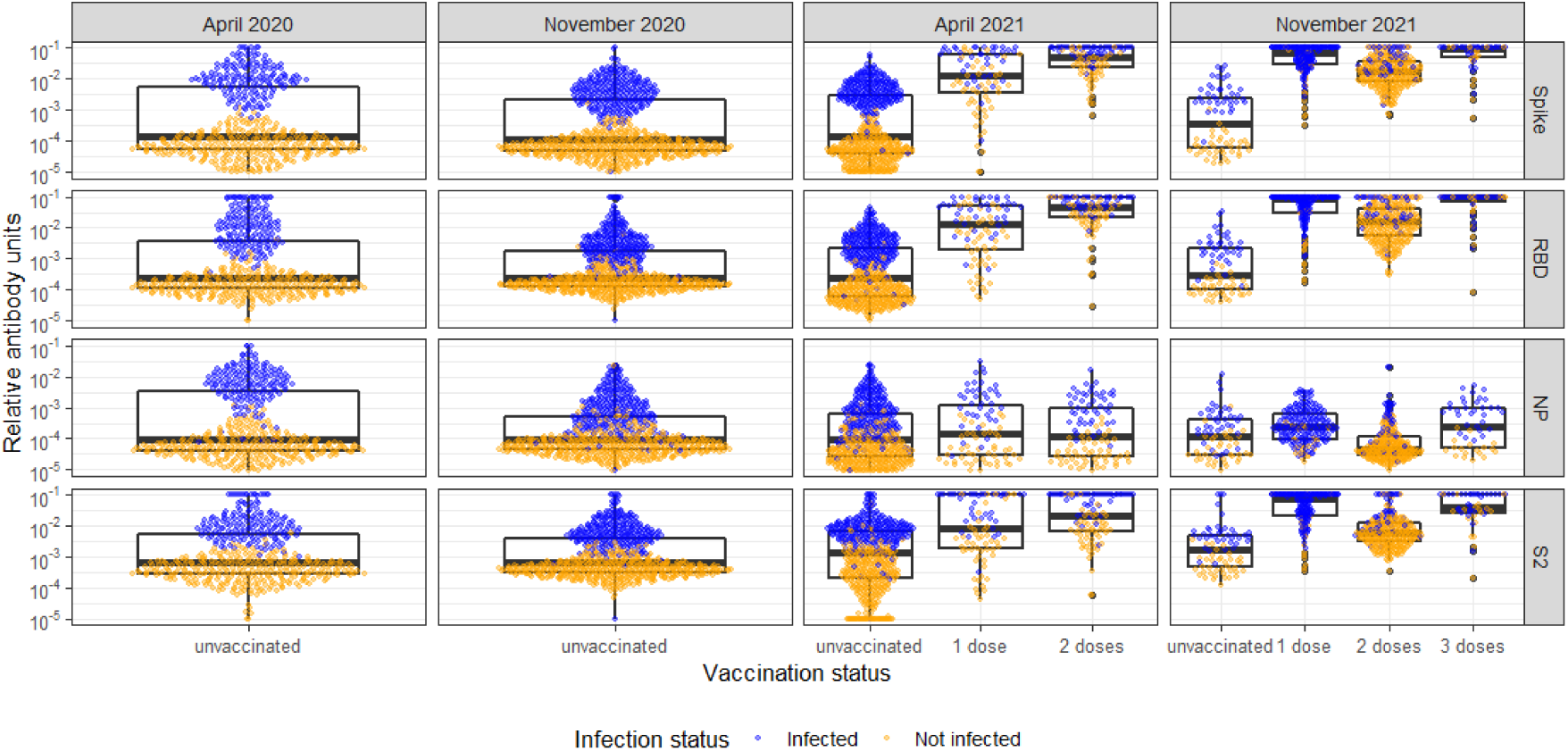
IgG antibody levels to the SARS-CoV-2 ancestral strain by vaccination status, moment of sampling and type of antigen. IgG antibodies are expressed as relative antibody units. Colours depict infection status.

Using the relative antibody units of Spike IgG, RBD IgG, S2 IgG, and RBD IgA, and the avidity index of IgG antibodies to Spike and RBD, as input for our random forest regression models, we translated these measurements into variant-specific neutralization activity. The estimated neutralization activity to the ancestral strain, Delta and Omicron are shown in Figure 4. Neutralization activity to the ancestral strain and Delta were derived with random forests regression models. Neutralization activity to Omicron was estimated by applying the 97.7% reduction relative to neutralization activity to the ancestral strain. Note that the neutralization activities are only shown for individuals who had immunity due to vaccination, infection or both.

**Figure 4.**
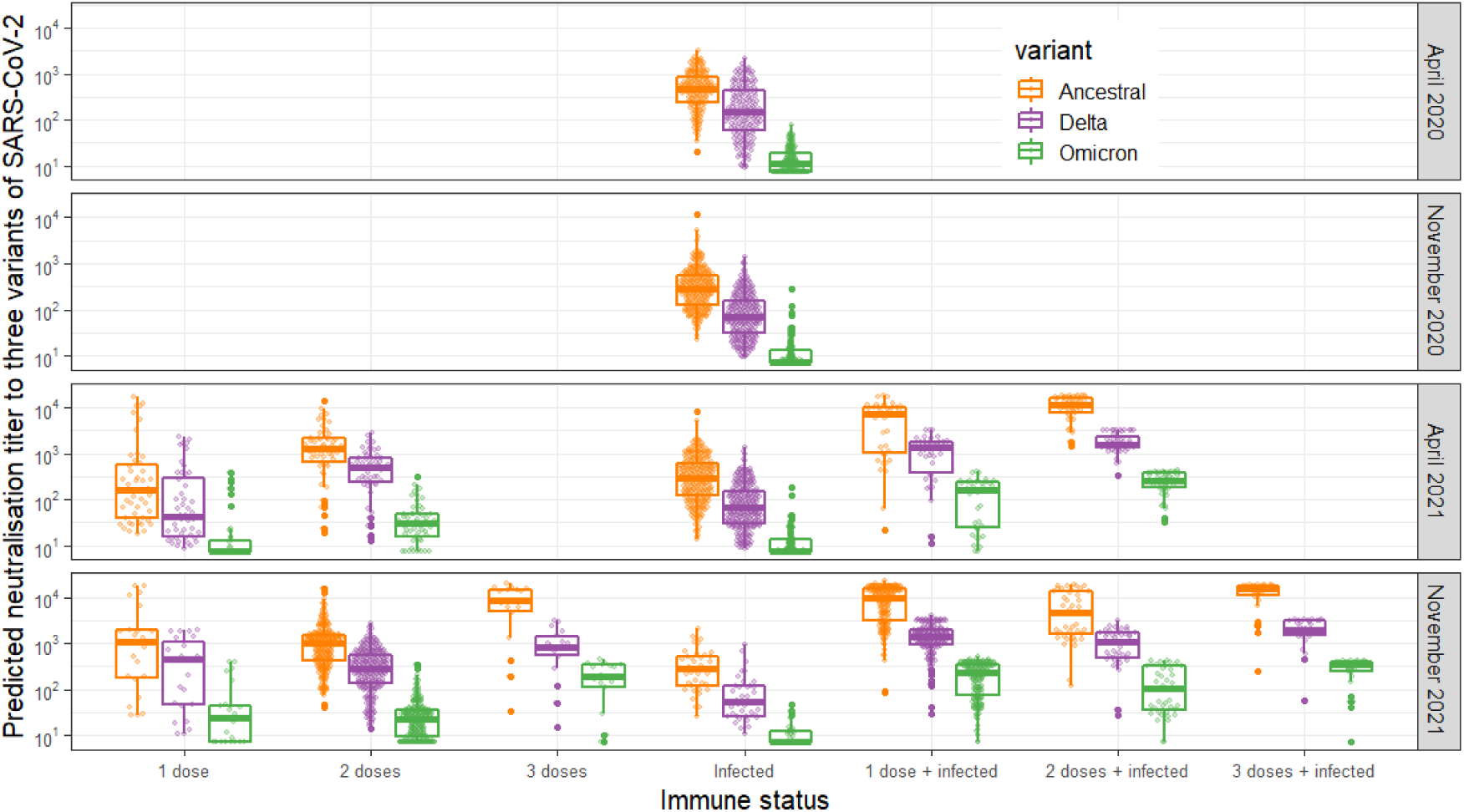
Predicted neutralization activity to three variants by immune status and date of sampling. Immune status was determined based on vaccinated status and infection status of the participants.

### Estimation of protection levels

Normalized neutralization titers were translated into estimates of protection. Estimates of protection were based on Khoury et al (7) using phase 2 immunogenicity and phase 3 efficacy estimates initially to describe the relationship between neutralization titers and protection. This was extended with vaccine effectiveness studies including real-world data and variant-specific protection by Cromer et al (12). These statistical relationships were not valid for Omicron, so we did not estimate protection against Omicron. Neutralization levels to Omicron drop to a level where there are unsufficient estimates of neutralization levels combined with vaccine efficacy estimates.

Relative to immunonaive individuals, the risk of COVID-19 and severe COVID-19 reduced with multiple vaccinations and/or past infection (Figure 5): the median reduced risk to COVID-19 caused by Delta was 42% among infected individuals and 96% among individuals vaccinated thrice. In line with the reduced neutralization titers to Delta vs ancestral strain, protection was lower against COVID-19 and severe COVID-19 due to Delta. Individuals with both vaccine-acquired and infection-acquired immunity had a further reduction in risk to COVID-19 or severe COVID-19. Amongst individuals with vaccine-acquired immunity only, the proportion of those under-protected against symptomatic COVID-19 (defined to be a reduced risk less than 50%) due to Delta was 35% after 1 dose, 14% after 2 doses, and 11% after 3 doses. Among individuals with both vaccine-acquired and infection-acquired immunity, the proportion of those under-protected against symptomatic COVID-19 due to Delta was 1% after 1 dose, 5% after 2 doses, and 3% after 3 doses.

**Figure 5.**
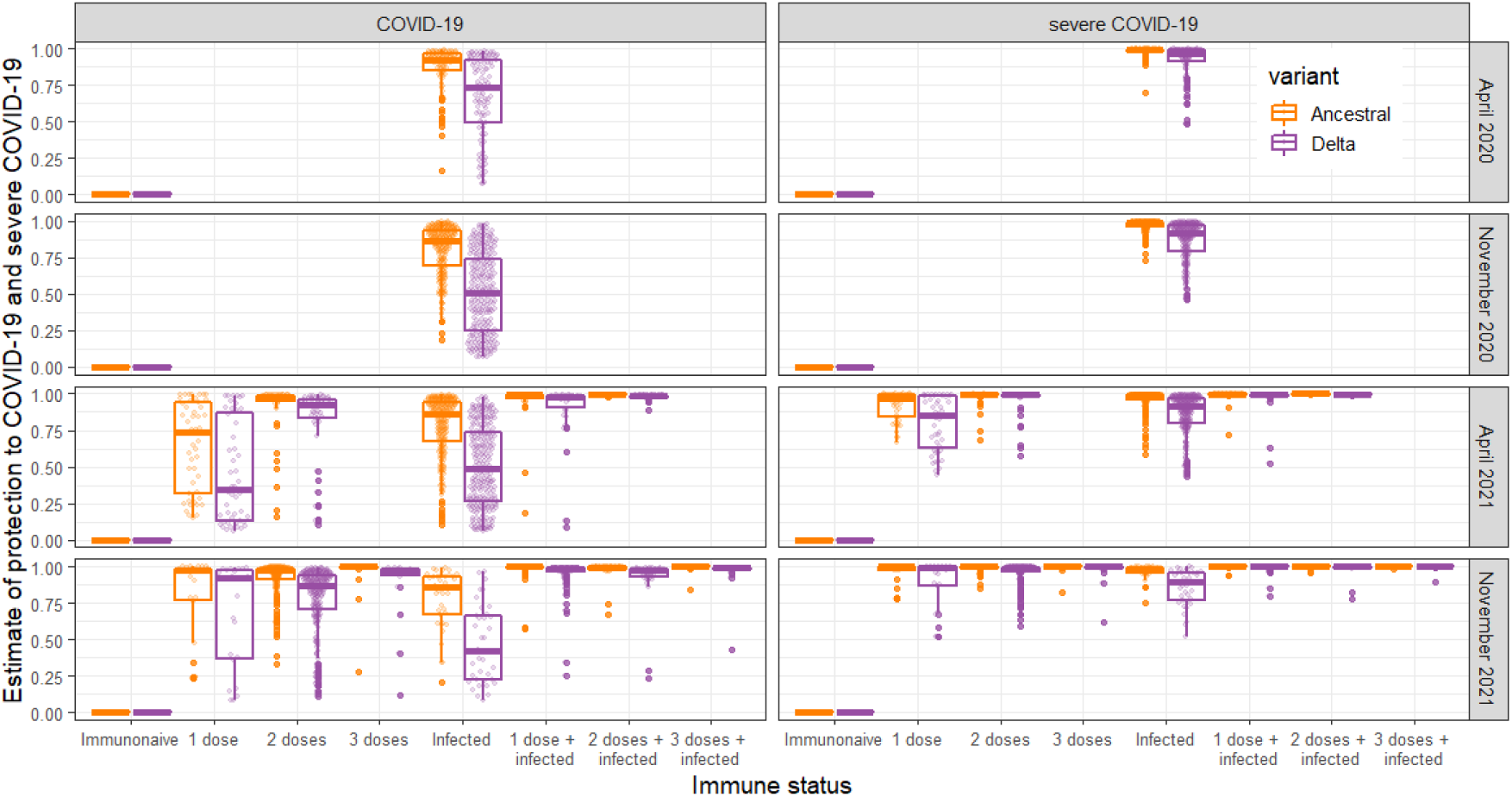
Estimates of reduced risk to COVID-19 and severe COVID-19 due to an infection with the ancestral and Delta variant among participants of the COVID-Oise cohort. Results are presented by sample collection sessions and by immune status due to vaccination and/or past infection.

By aggregating individual protection estimates, we estimated the susceptibility to COVID-19 at the population level. We grouped the COVID-Oise cohort by age group, immune status (a combination of vaccination status and infection status), and summarized protection by identifying the median protection for each of these aggregated groups. The stacked protection by age group revealed that the oldest age group had the highest reduced risk of COVID-19, partly caused by the high vaccination coverage that was ahead of other age groups (Figure 6). In Figure 6, we visualize the population-level protection for the COVID-Oise cohort in December 2021. These findings can be extrapolated to provide an assessment of population-level immunity in the rest of France in December 2021. Based on data on vaccine doses collated by Santé Publique France and reported infections adjusting for underreporting (20), the immune profile of Crépy-en-Valois is representative of the rest of France (Figure S10).

**Figure 6.**
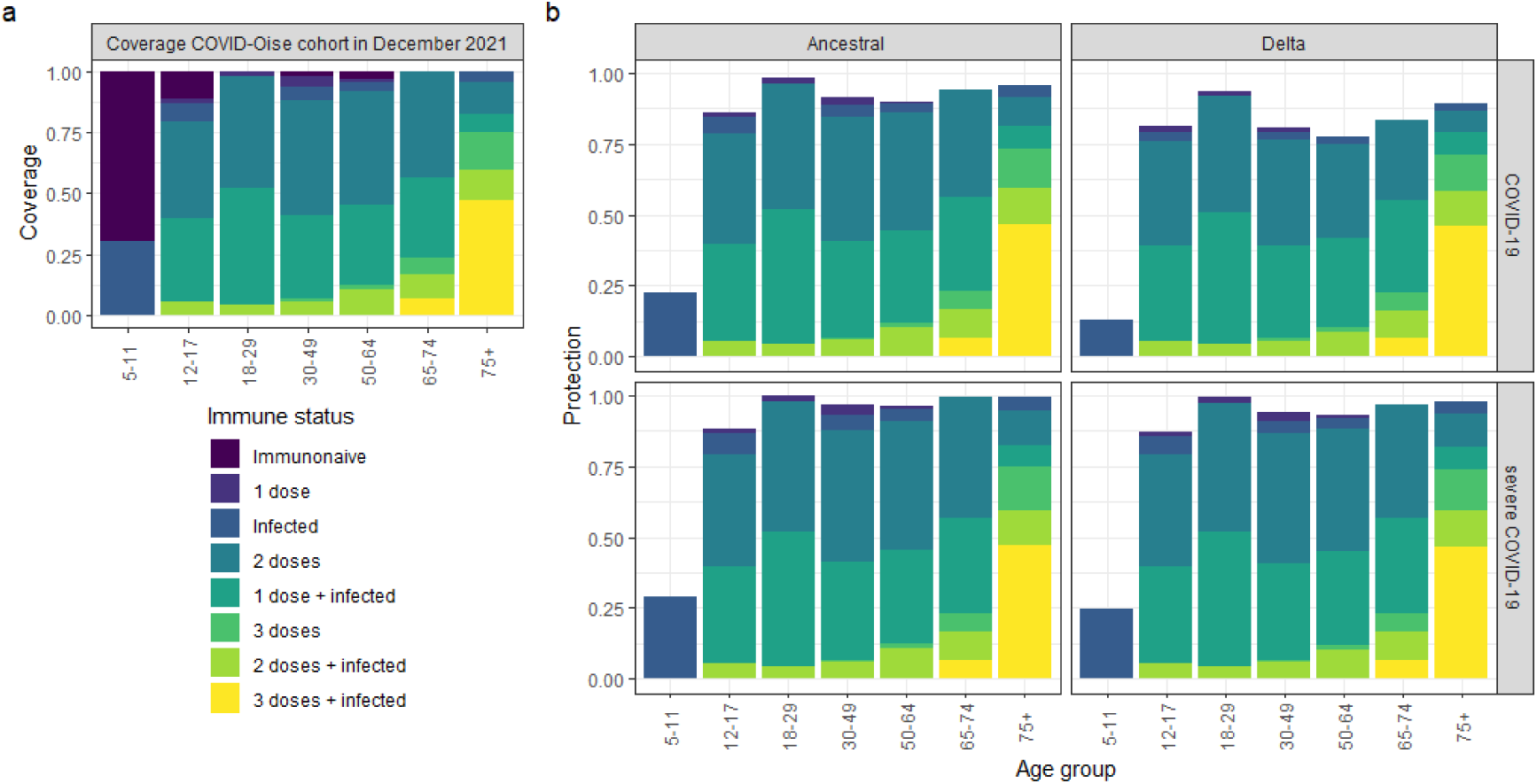
Aggregated protection against COVID-19 and severe COVID-19 in December 2021. (a) Coverage of vaccination and infection status by age group in participants of COVID-Oise cohort in December 2021. (b) Stacked median reduced risks to COVID-19 and severe COVID-19 by variant and age group.

## Discussion

Seroprevalence studies are widely used to provide an indication of the amount of transmission and vaccination coverage in a population. Immunity resulting from infection or vaccination does not guarantee prevention of infection, illness, or hospitalization by SARS-CoV-2. Our novel method allows seroprevalence estimates to be translated into estimates of protection. Using high-throughput multiplex assays with variant-specific antibodies allows us to identify under-protected individuals who may be targeted with additional vaccine doses.

Our estimates are in line with observed vaccine effectiveness estimates for both the ancestral and Delta strains (Figure S11). We estimated protection levels by time since vaccination, which were comparable in a series of observational and randomized controlled trials of vaccines (21-27). We observed increased protection for individuals with hybrid immunity in line with others (28, 29). Individuals with one vaccination and a confirmed infection were better protected than individuals with two doses.

Multiple lines of evidence demonstrate that neutralizing titers are associated with protection against both symptomatic and severe COVID-19 (7-9), but T-cell mediated immunity is also known to play a critical role (30). As many T-cell epitopes are not mutated in variants of concern, the contribution of T cells to protective immunity is likely to remain, most notably for protection against severe COVID-19. A limitation of our study was that we assessed levels of immunity from serum only. There is clearly also a role for mucosal immunity in protecting against SARS-CoV-2 infection, especially in the case of infection-acquired immunity. For the last three session in our study we collected nasopharyngeal samples, which we plan to incorporate in future research.

Our analysis is dependent on the suitability of neutralizing titers as a correlate of protection against symptomatic COVID-19, based on meta-analyses of vaccine studies (7, 8). This assumption is supported by an analysis of data from phase 3 trials of Moderna’s mRNA-1273 vaccine, which indicated that 68% of vaccine efficacy can be explained by neutralizing titers (31). This leaves up to 32% variation that may be explained by other effects such as cellular immunity or host factors. An additional limitation is that the evidence base for neutralizing titers as a correlate of protection is built on studies of infection with the Ancestral variant. However, antibody levels have been observed to be associated with reduced infection with other variants, most notably Delta (32). Although neutralizing titers have frequently been shown to be associated with protection against severe COVID-19 (7-9), there is a weaker evidence base for their use as a correlate of protection. A final, important limitation is that there is uncertainty in the statistical relationships utilized in this analysis. When considering the inferred protection from symptomatic COVID-19 obtained by analyzing a sample, there will be substantial uncertainty in that individual’s estimated protection. This uncertainty will limit the use of our methods for diagnosing under-protected individuals. However, in this study we focus on aggregated protection across large numbers of samples, where the effects of uncertainty are diminished, but not removed.

Our final cross-sectional analysis of population-level protection is from December 2021. At this time, many individuals had recently received their second or third vaccine doses, and consequently had high antibody responses. In addition, many individuals had prior SARS-CoV-2 infection resulting in stronger immune responses. Furthermore, there are interactions between numbers of vaccine doses and infection status, most notably as a consequence of the policy recommendation in 2021 to consider individuals with documented previous infection fully immunized after only one vaccine dose (and eligible for the French “passe sanitaire”). (Figure 3, November 2021).

The December 2021 cross-section occurred just before the emergence of the Omicron variant in France, replacing the previously dominant Delta variant. By accounting for the 97% reduction in neutralization of Omicron compared to the Ancestral variant, we were able to indirectly estimate Omicron neutralization titers. However, we did not attempt to infer protection against Omicron infection due to a lack of a validated correlate of protection. As a substantial proportion of the French population has been infected by Omicron variants since December 2021, our estimates are not representative of the current immunity present in the French population. However, with the inclusion of antigens from Omicron or any other relevant variant that may appear in future, our assay can be readily extended and applied.

## Data Availability

All data produced in the present study are available upon reasonable request to the authors.

## Acknowledgements

The authors wish to thank the participants who agreed to participate into the different studies and the medical and paramedical teams who were involved in sample and data collection. We thank the teams of Crépy-en-Valois town hall and the Director and technical services of the local hospital for their help in the implementation of the COVID-Oise study. We thank the ICAReB technical team for management and distribution of the samples. We would like to acknowledge Devan Sinha for proposing the idea of the immunity wall to COVID-19

## Author contributions

LP, MNU, SFP, OS, AF and MTW planned and designed the study. LP, SP, MA, CD, ADK, SC organised enrolment and follow-up of study participants and processed the samples together with LG, LT, and FD. DP and TB assessed neutralization activity of samples with the neutralization assay. LG, SP, FD, MA performed tests with the bead-based multiplex assay, and MA with the LuLISA. TW, LP, LT, ADT, MW performed the data analysis.TW wrote the first draft of the manuscript. All authors contributed to critically revising the manuscript and approved the final version. Project administration was managed and supervised by MNU, SFP, OS, AF, and MW.

## Data availability

All data produced in the present study are available upon reasonable request to the authors.

