## Supplementary figures and table for "Humoral Immunity to SARS-CoV-2 and Inferred Protection from Infection in a French Longitudinal Community Cohort"

**Supplementary material**

| **Table S1. Characteristics of participants of whom serum samples were obtained to correlate relative antibody units and neutralization activity from the virus neutralization studies.** | | | |
| --- | --- | --- | --- |
| Characteristic | Sub-group | N | % |
| Sex | Female | 90 | 45 |
|  | Male | 108 | 54 |
|  | Missing | 106 |  |
| Age (median, range) |  | 56 | 42 - 60 |
| Immunity status | Infection-acquired | 106 | 30 |
|  | AstraZeneca – one dose | 11 | 4 |
|  | AstraZeneca – two doses | 20 | 7 |
|  | Jansen – one dose | 15 | 5 |
|  | Pfizer – two doses | 89 | 29 |
|  | Sputnik – two doses | 34 | 11 |

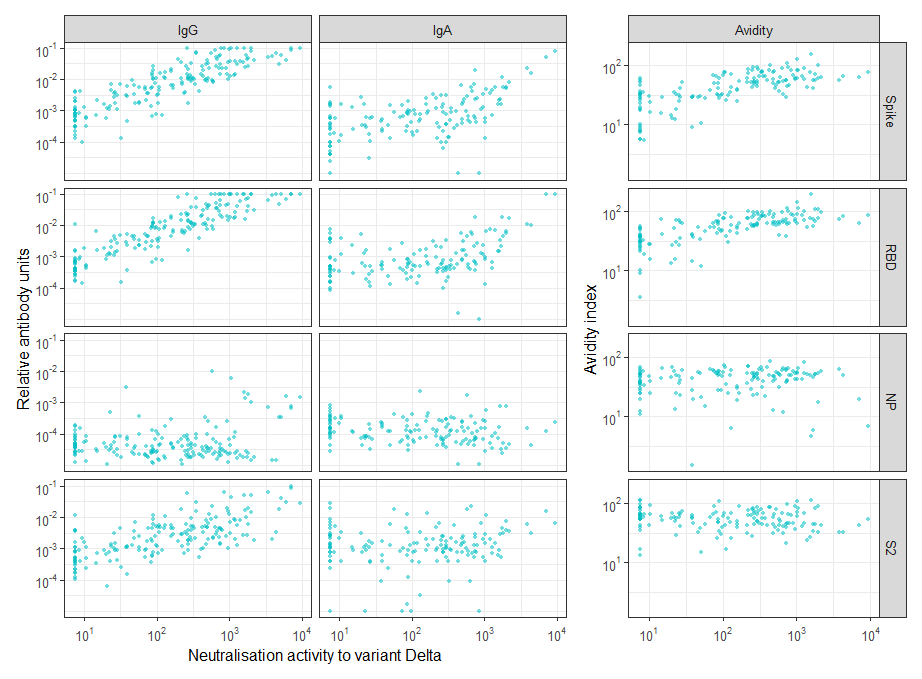

Figure S1. Correlation between relative antibody units and avidity index of four antigens with neutralization activity to variant Delta.

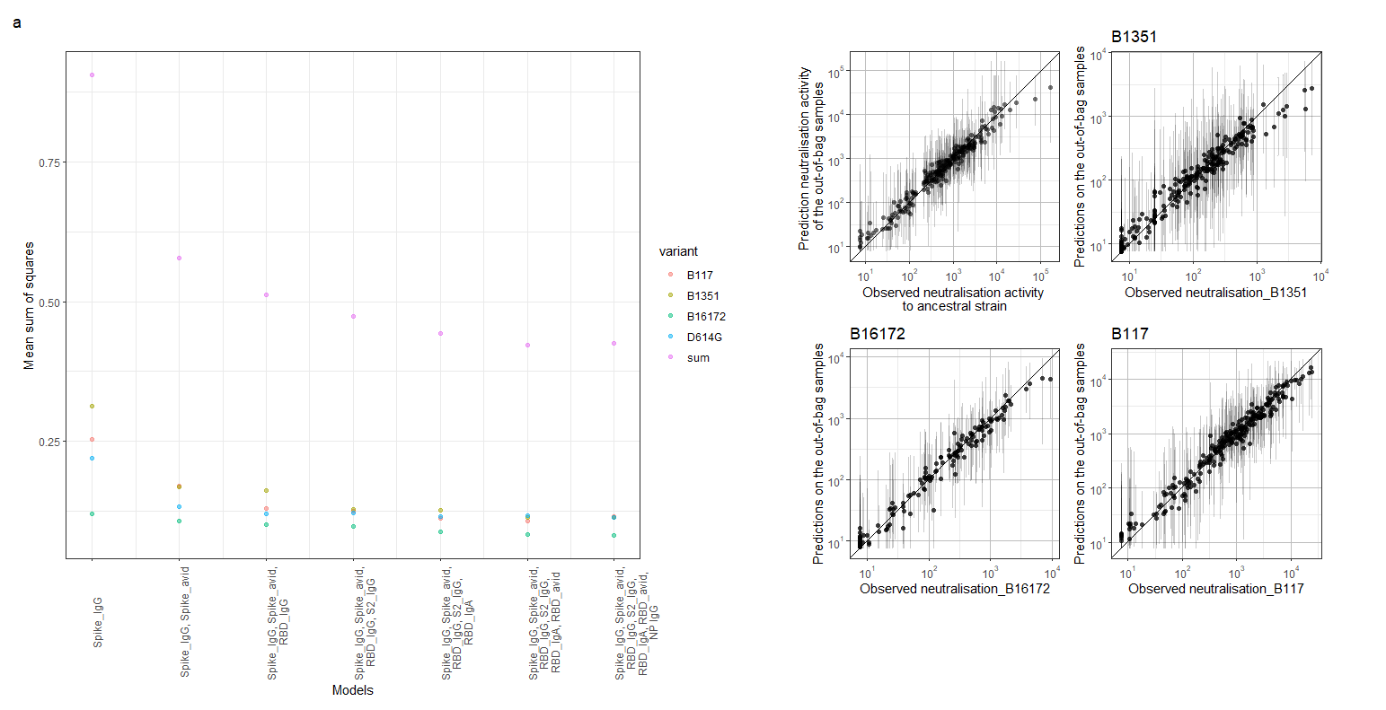

Figure S2. Development of a random forest regression model predicting neutralizing activity. (a) Basic regression model contained Spike IgG only. This model was complemented with the antigen producing the largest drop in the mean sum of squares for four models predicting neutralising activity to four strains. After model 6, no further reduction was observed. (b) These are the four random forest regression models predicting neutralising activity to D614G, B1351, B16172, and B117. The points reflect the means of the prediction of the out-of-bag samples. The vertical lines show the 2.5 and 97.5 percentile of the predictions.

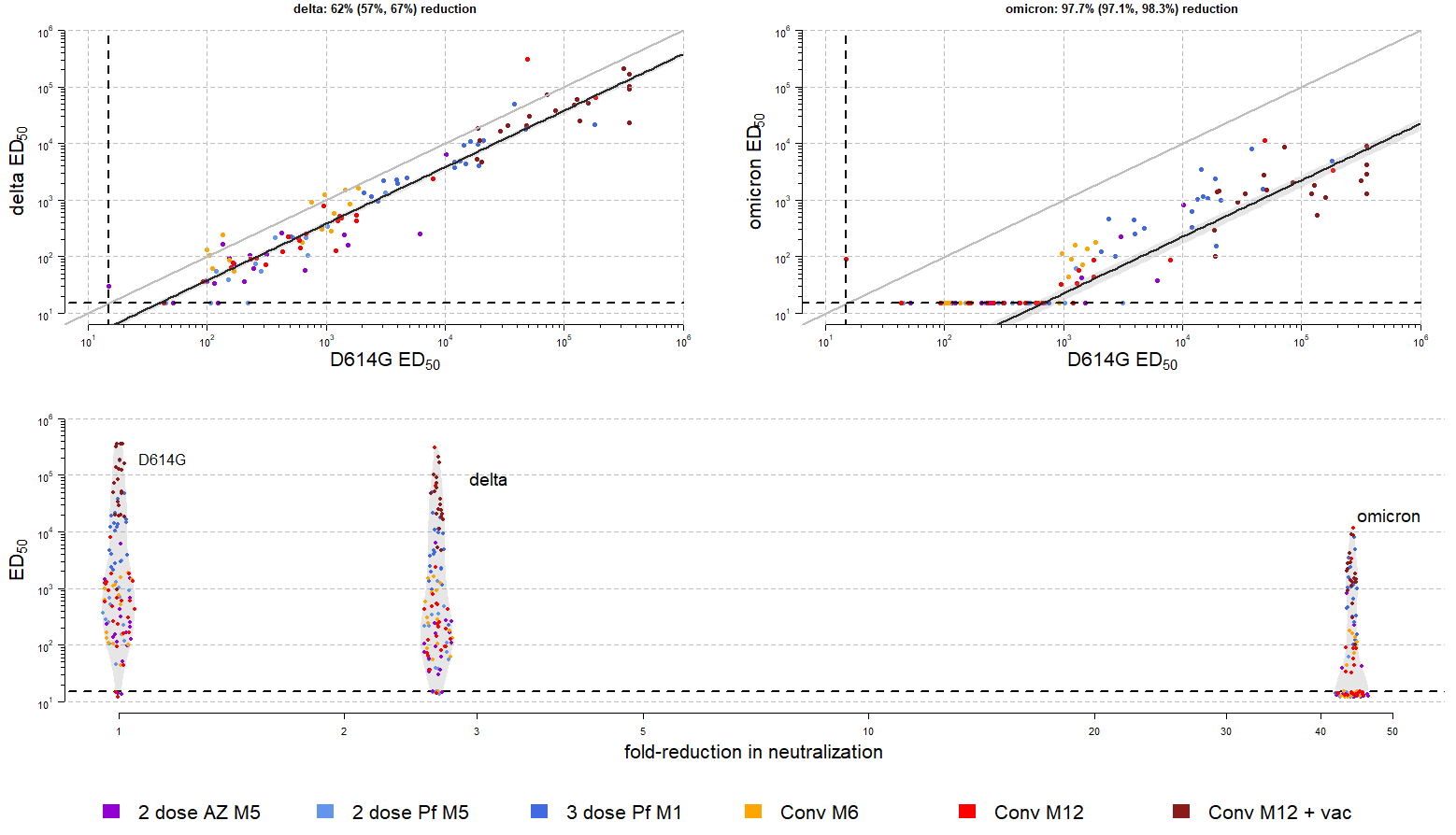

Figure S3. **Neutralization activity decreased almost 3-fold between ancestral strain and Delta, and more than 40-fold between ancestral variant and Omicron.** In two panels at the top, the fit of the censored linear regression is shown. In the figure in the lowest panel, this is converted into a fo*ld-reduction in neutralisation.*

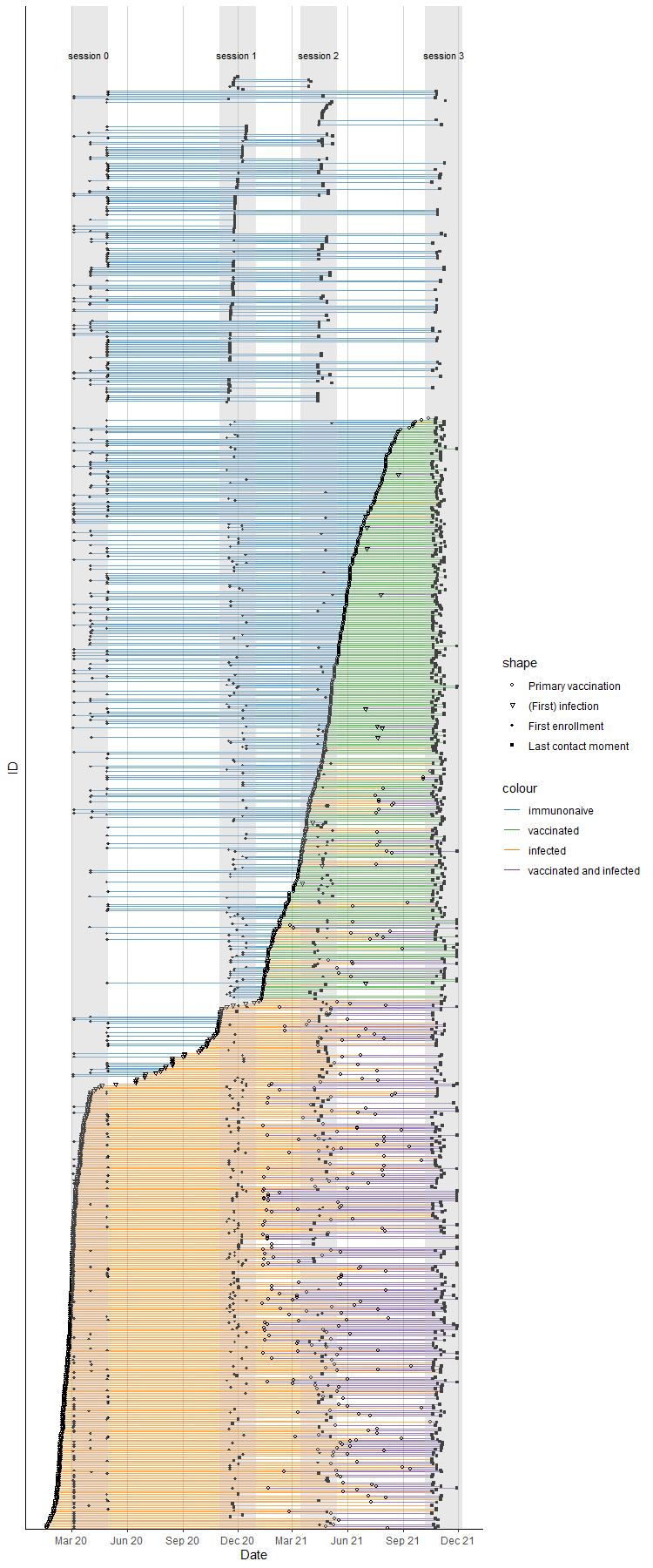
Figure S4. **A complete visualization of all events in our study, including infections, vaccinations and enrolment activity in the longitudinal cohort study.** Colors depict the status of the participants in the study, which included immunonaive, vaccinated, infected, and infected and vaccinated.

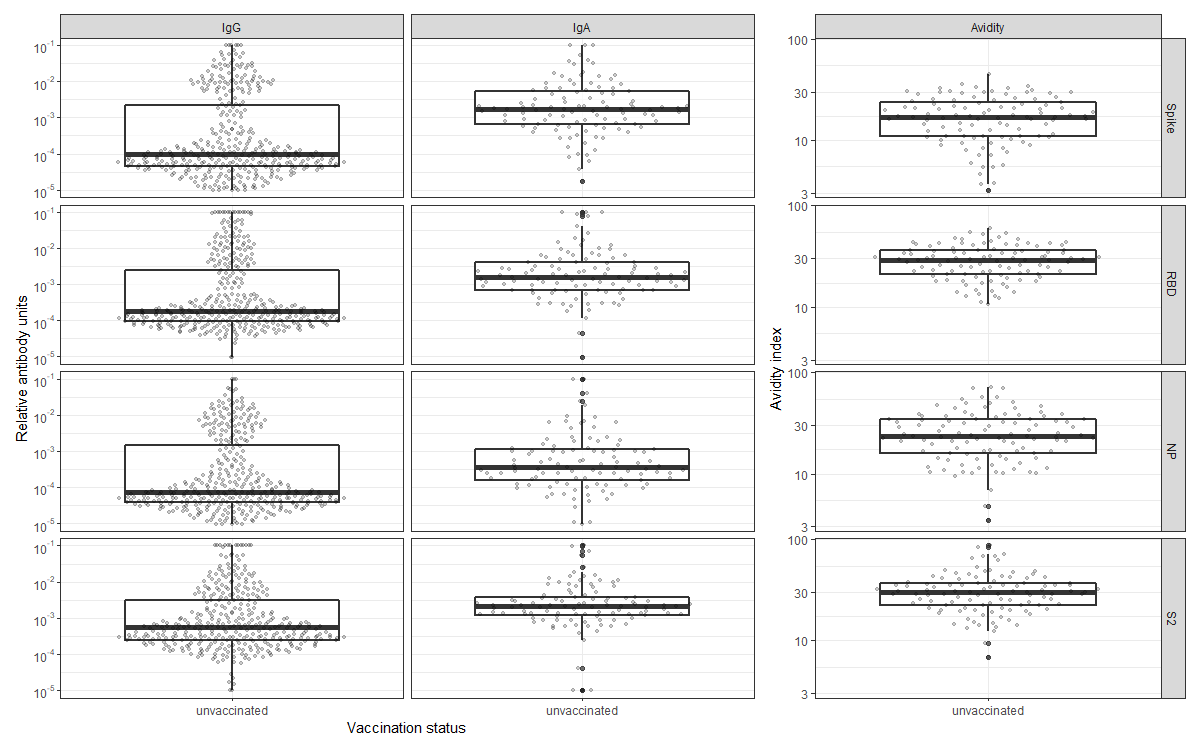

Figure S5. **Antibody distributions by vaccination status, isotype and antigen in a cohort sampled around April 2020.**

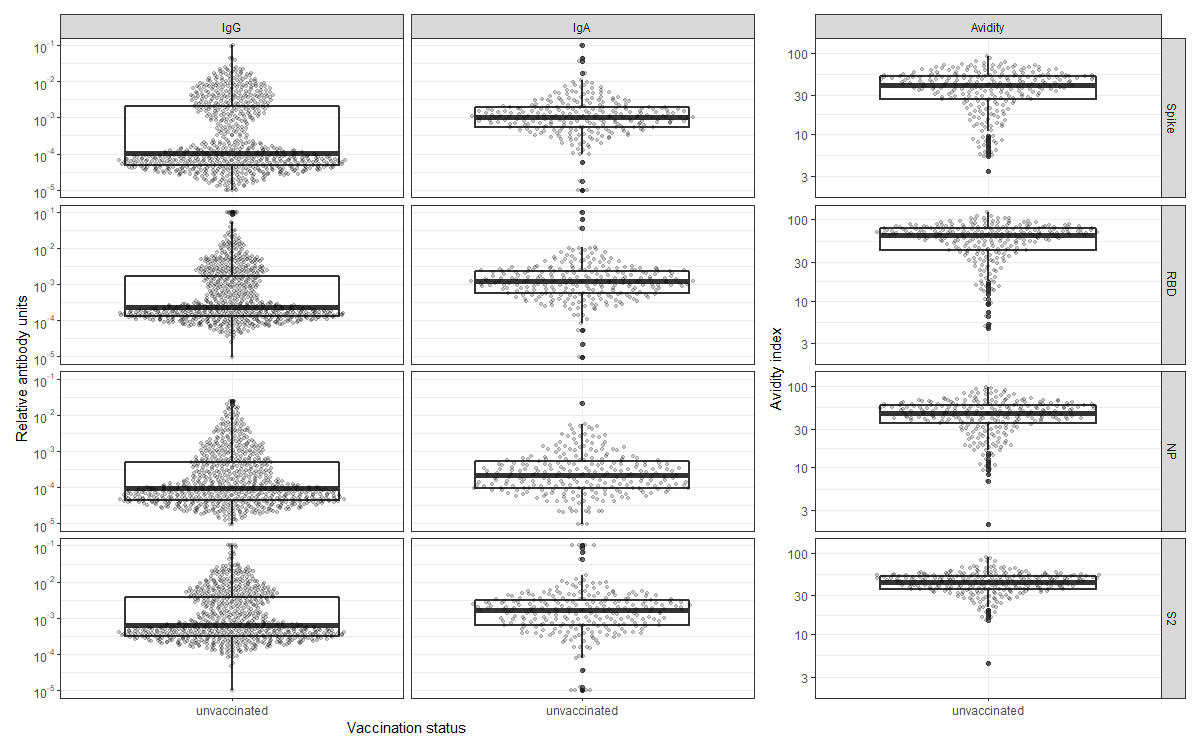

Figure S6. **Antibody distributions by vaccination status, isotype and antigen in a cohort sampled around November 2020**

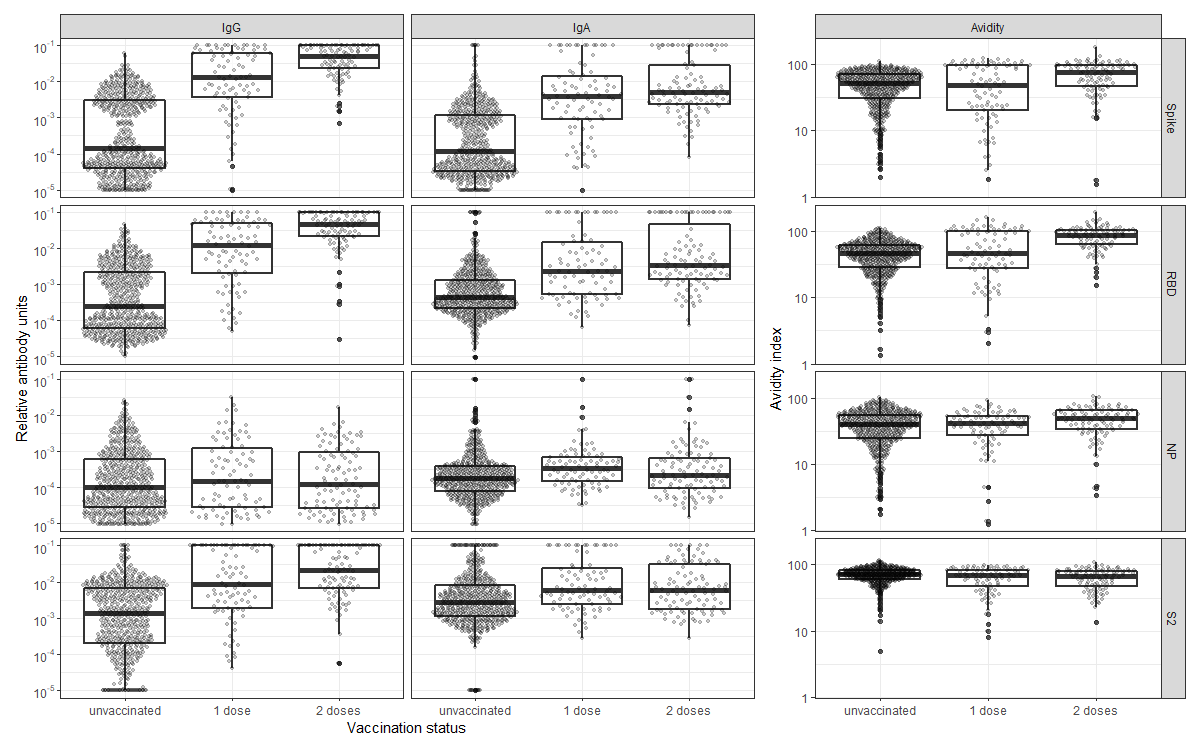

Figure S7. **Antibody distributions by vaccination status, isotype and antigen in a cohort sampled around April 2021**

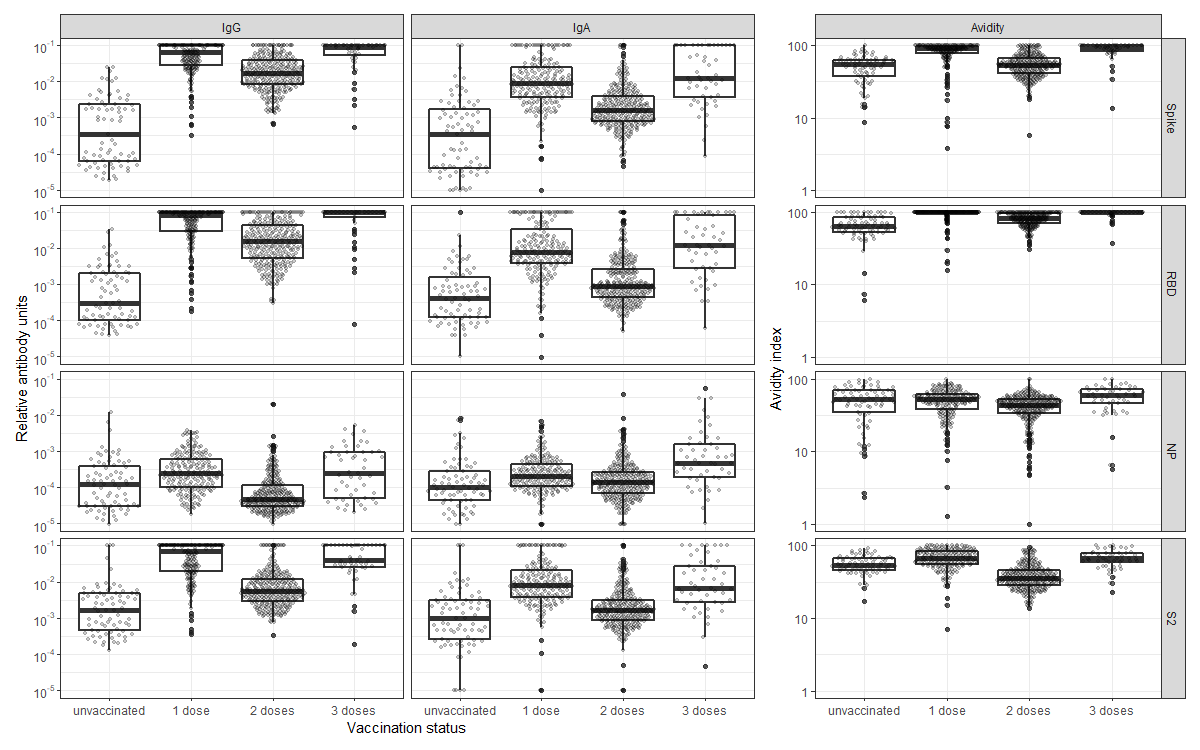

Figure S8. **Antibody distributions by vaccination status, isotype and antigen in a cohort sampled November and December 2021.**

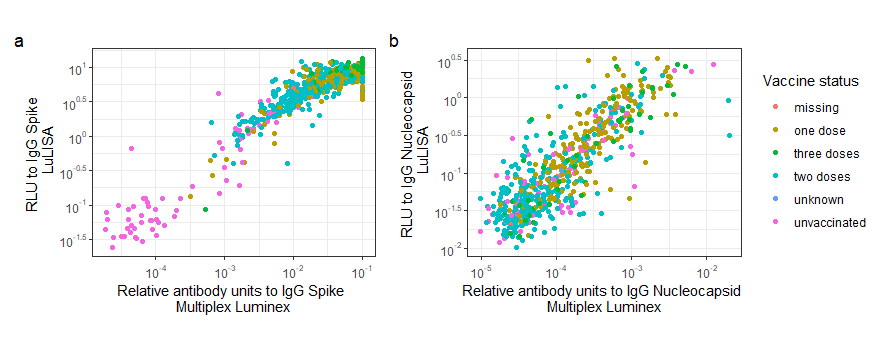

Figure S9. **Measurements of antibodies to Spike (a) and Nucleocapsid (b) correlated well between the multiplex Luminex assay and the LuLISA assay.** A clear separation was observed between vaccinated and unvaccinated individuals. The samples shown here are from the COVID-Oise cohort in sampled in November 2021.

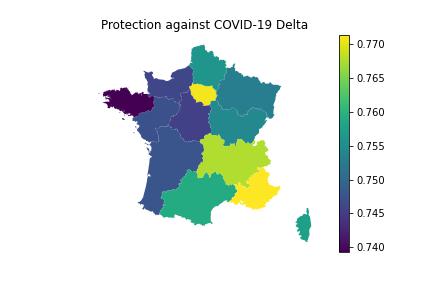

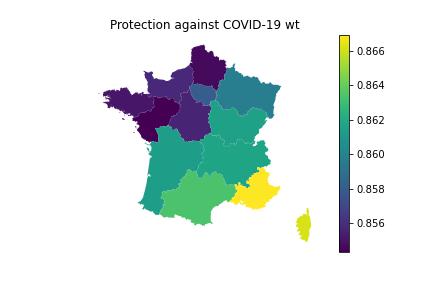

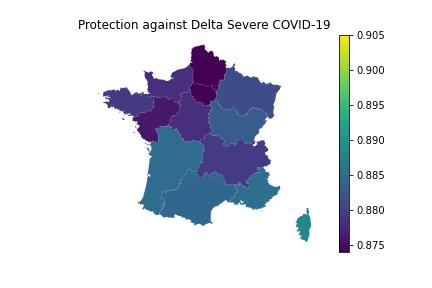

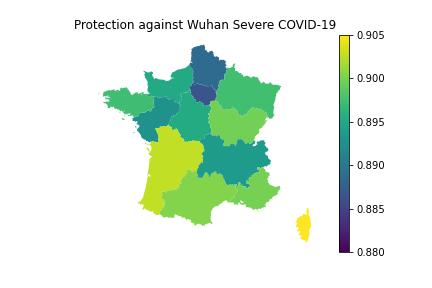

Figure S10. Protection estimates to COVID-19 and severe COVID-19 caused by the ancestral variant or Delta variant, by region. Protection estimates are aggregated estimates by age and dependent on immunity status. Vaccination status by age group were derived from Santé Publique France and infection status is derived in a similar manner as stated in Hozé et al (20).

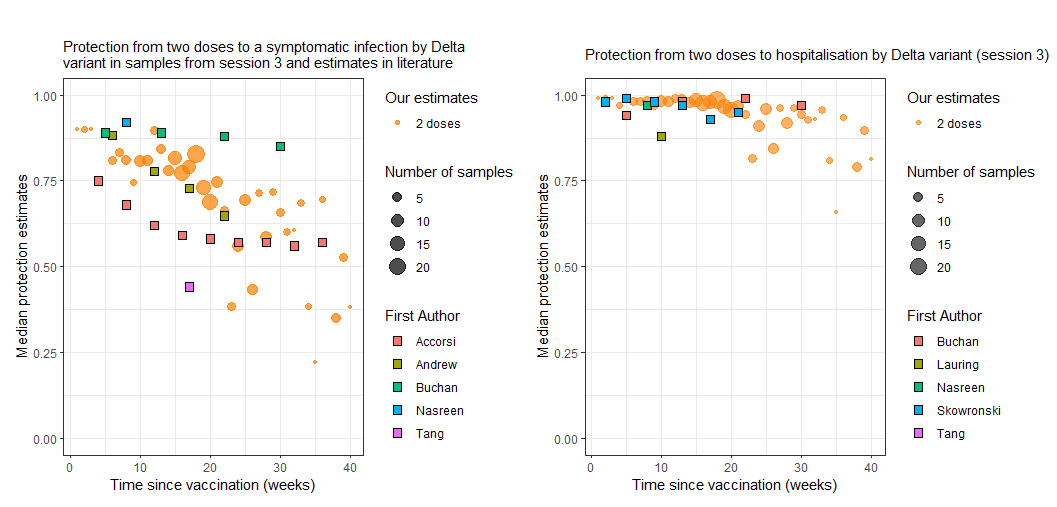

Figure S11. Estimated protection estimates and vaccine effectiveness estimates from literature correlate well. Our estimates by time since vaccination overlap with vaccine effectiveness estimates from the field. Vaccine effectiveness estimates are from(22-27).

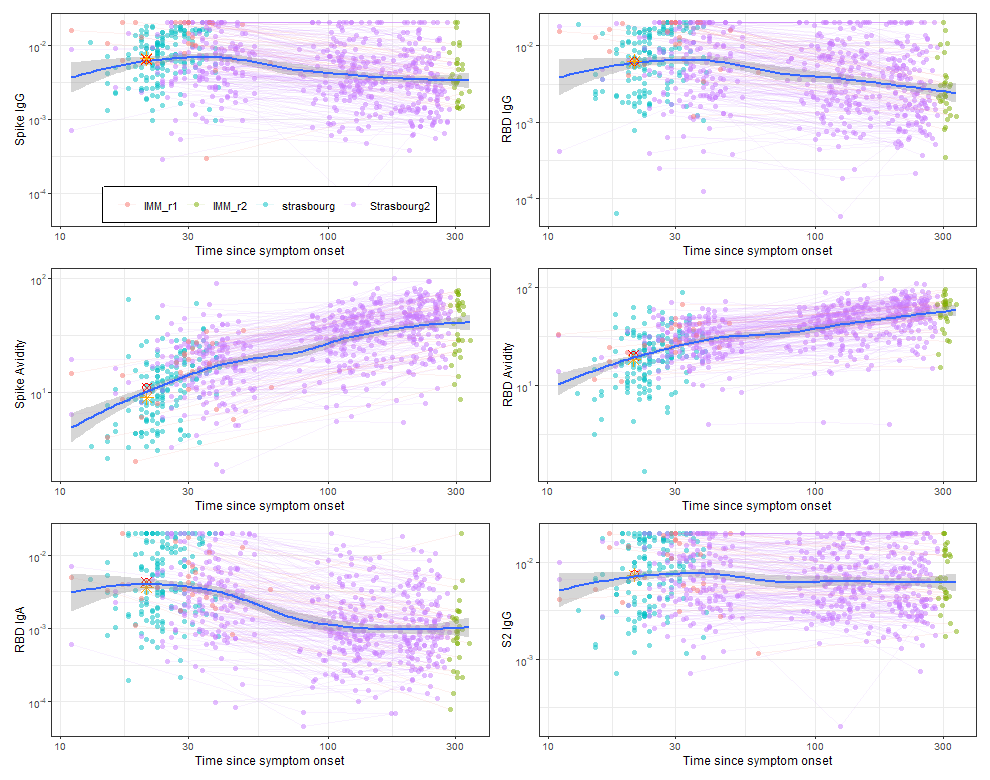

Figure S10. **Estimation of convalescent serum.**
